# The ambulatory arterial stiffness index is not a measure of arterial stiffness in childhood

**DOI:** 10.1101/2025.10.24.25338706

**Authors:** Plamen Bokov, Elodie Surget, Cherine Benzouid, Benjamin Dudoignon, Julien Hogan, Christophe Delclaux

## Abstract

**Background:** The ambulatory arterial stiffness index (AASI) has emerged as an ambulatory blood pressure monitoring (ABPM) measure of stiffness and is supposedly useful in younger subjects. The objective of our cross-sectional study was to evaluate the relationships between the AASI and indices of arterial stiffness in a pediatric population at risk of hypertension.

**Methods:** This was a cross-sectional study of children/adolescents (8–18 years) whose pulse wave velocity (PWV: carotid-to-femoral cf-PWV and heart-finger hf-PWV), augmentation index (AIx; normalized at 75 bpm: AIx_75_), systemic arterial stiffness (aortic pulse pressure/stroke volume, measured via pulse contour analysis) and ABPM were measured on the same day. At-risk populations were vascular remodeling (preterm birth, n=44 and chronic kidney diseases, n=7) and hyperkinetic causes (congenital central hypoventilation syndrome, n=14 and psychostimulant treatment, n=10).

**Results:** The mean age of the 75 participants was 12.3 ± 2.5 years (34 girls), and their mean AASI was 0.33 ± 0.17. AASI did not correlate with cf-PWV, hf-PWV, AIx or systemic arterial stiffness. In contrast, the AASI significantly correlated with both systolic and diastolic BP at night (R= -0.23; p=0.048 and R= -0.33; p=0.004, respectively). Systemic arterial stiffness correlated with hf-PWV and AIx_75_ (R= 0.35; p=0.004 and R= -0.34; p=0.013, respectively). Based on ABPM, 15/75 (20%) participants had hypertension, and they had higher cf-PWV than participants without hypertension (5.64 ± 0.70 vs 4.92 ± 0.78 m/s, p=0.002) and not different AASI values (0.34 ± 0.14 vs 0.32 ± 0.18, p=0.756).

**Conclusion:** AASI is not a measure of arterial stiffness in childhood.

## Introduction

The ambulatory arterial stiffness index (AASI) has emerged as an increasingly important and novel ambulatory blood pressure monitoring (ABPM) measure.^1^ It is supposed to be distinct from the majority of other ABPM measures in that it is both a measure of BP variability and an indirect measure of arterial stiffness.^2^ It is calculated as 1-minus the regression slope of ambulatory diastolic versus systolic BP.^2^ In stiffer arterial trees, the systolic-diastolic BP regression slope tends to be lower near 0, and the AASI is greater (closer to 1). The AASI has been shown to vary considerably in cases where 24-h ambulatory blood pressures and pulse pressures remain similar. AASI is correlated with both pulse wave velocity and the arterial augmentation index in adults,^2,3^ as are subclinical markers of target organ damage, including carotid intimal thickness, left ventricular hypertrophy and worsening renal function.^4^ Furthermore, Simonetti *et al*. reported that hypertensive children had higher AASI values than their normotensive healthy counterparts did,^5^ suggesting its usefulness in childhood.

Nevertheless, Schillaci *et al*. reported that the AASI is strongly dependent on the degree of nocturnal BP decrease in hypertensive patients and that the relationship between the AASI and a widely accepted measure of aortic stiffness, such as pulse wave velocity, is weak and, importantly, affected by other factors.^3^

Overall, whether the AASI, despite its prognostic value,^4^ is truly a marker of arterial stiffness remains debated in adult patients and has not been evaluated in childhood, a period in which hypertension is often due to hyperkinetic causes (due to sympathetic overactivity) rather than vascular remodeling processes, which are long-term consequences of hypertension.^6^ Thus, the main objective of our study was to assess the correlation between the AASI and the aortic pulse wave velocity (the gold standard of arterial stiffness^7^), which is the speed of wave travel that intrinsically represents arterial stiffness, according to the Bramwell‒Hill formula, and to assess whether hypertensive children already have increased AASI. Our secondary objectives were to evaluate the relationships between the arterial stiffness indices studied since they have not been determined in childhood and to compare subjects according to the presence of hypertension and according to the underlying cause (hyperkinetic versus vascular).

## Materials and methods

### Design and Patients

This was a cross-sectional study of children/adolescents (8--18 years old) referred for cardiovascular disease prevention (subjects at risk of hypertension) who had pulse wave velocity (PWV) measurements and ambulatory blood pressure monitoring (ABPM) on the same day (single-day hospitalization). The following four conditions were included: preterm birth, which has been associated with increased hypertension prevalence;^8^ treatment with a psychostimulant (methylphenidate) for narcolepsy type 1, which has also been associated with increased BP;^9^ CCHS, which is associated with neurogenic hypertension;^10^ and chronic kidney disease (CKD, i.e., autosomal dominant polycystic kidney disease or CKD on hemodialysis), which is the most frequent cause of secondary hypertension in childhood.^11^ Among these four conditions, two have been associated with vascular remodeling (preterm birth and CKD^12–14)^, whereas the other two conditions have hyperkinetic causes (CCHS and psychostimulant treatment). The only noninclusion criterion was a nonreliable ABPM defined by a percentage of successful measures < 70% of all attempted readings.^15^ Our local ethics committee approved this study (PHENOPA: N° 2024-199). The subjects and their parents were informed of the prospective collection of their data for research purposes, and they could request that they be exempted from this study in accordance with French law on noninterventional observational research. This study complied with the STROBE criteria for cross-sectional studies.

### ABPM

Ambulatory BP (nondominant arm) was recorded via a validated oscillometric device (Spacelabs model 90207, SpaceLabs, Inc.) set to take a reading every 15 minutes throughout the day (6:00 AM to 10:00 PM) and every 30 minutes throughout the night (10:00 PM to 6:00 AM), according to recommendations.^15^ Normal daily activities were allowed and encouraged. Day and night subperiods were defined according to patients’ diaries. From individual 24-hour recordings, the device calculated the regression slope of diastolic BP on systolic BP values. The AASI was calculated as 1 minus the regression slope, as proposed by Li *et al*.^2^

ABPM classification defines hypertension in children (< 13 years) as daytime or nighttime SBP or DBP ≥ 95^th^ percentile and in adolescents (≥ 13 years) as daytime BP ≥ 130/80 mmHg or nighttime BP ≥ 110/65 mmHg.^15^ Masked hypertension was defined as hypertension during ABPM when BP was within the normal range in our laboratory (at the office).

### Pulse wave velocity (PWV) and tonometry

Aortic (carotid-to-femoral) cf-PWV was calculated from measurements of common carotid and femoral artery wave forms via an automatic applanation tonometry-based device, the SphygmoCor Vx system (AtCor), in the supine position after 10 min of rest. The direct straight distance between the two measurement sites was measured via a tape measure, and the cf-PWV was then calculated (distance × 0.8^16^). The aortic pressure waveform was deduced from the common carotid artery waveform,^7^ allowing the recording of the aortic SBP, aortic pulse pressure, subendocardial viability ratio and augmentation index (AIx) to further normalize to a heart rate of 75 bpm (AIx_75_). We further calculated the central-to-peripheral amplification ratio (aortic SBP/SBP), which characterizes the stiffness gradient.

### Heart rate variability (HRV) and baroreflex sensitivity (BRS) measurements

We continuously monitored ECG, noninvasive arterial pressure (using the vascular unloading technique and a CNAP® Monitor from CNSystems, Austria), and respiratory activity (using a piezoelectric belt from BIOPAC Systems, France) and digitized signals with the MP-160 system (BIOPAC Systems) as previously described.^17^

Using ECG and noninvasive arterial pressure signals, heart-finger hf-PWV was calculated as described by others.^18^ The direct distance between the sternal notch and fingers was measured. The ratio of hf-PWV/cf-PWV was calculated since it also characterizes the stiffness gradient.

A beat-by-beat data series taken during 5 minutes of rest in the supine position was selected offline. The baroreflex was analyzed with AcqKnowledge’s baroreflex sensitivity analysis (BIOPAC Systems, Paris, France) software package via the sequence method. We calculated the means of the ascendant and descendant BRSs, as previously described.^17^ HRV was analyzed with HRVanalysis software 1.1 downloaded from https://anslabtools.univ-st-etienne.fr and validated by Pichot *et al*.’s method,^19^ as previously described.^10,17^ After the fast Fourier transform, the power spectrum indices were calculated. Very low frequencies (VLF: 0–0.04 Hz), low frequencies (LF: 0.04–0.15 Hz), and high frequencies (HF: 0.15–0.40 Hz) were calculated. LF and HF were also expressed as normalized values, LFnu and HFnu.

CNAP® Technology uses pulse contour analysis applied to the CNAP® blood pressure waveform to provide full hemodynamics, including stroke volume, cardiac output and systemic vascular resistance (SVR). Cardiac output measurement via CNAP® technology has been validated against invasive clinical standards (i.e., thermodilution) and has shown high agreement in terms of accuracy and trending ability.^20^

Systemic arterial stiffness was calculated via the following formula: aortic pulse pressure/stroke volume,^7^ expressed as mmHg/mL.

### Statistical analyses

Sample size calculation: Based on the methods of Schillaci *et al*. and Li *et al*., the correlation coefficient between the AASI and PWV ranges from 0.28 to 0.51.^2,3^ The total sample size required to determine whether a correlation coefficient of 0.40 (mean value between 0.28 and 0.51) differs from zero, with a type I error rate of 0.05 and a type II error rate of 0.80, was 47. This sample size was increased to 75 to allow multivariate analyses with five variables (15 subjects per variable), the number of variables previously used.^2,3^ Continuous variables were tested to detect substantial deviations from normality by computing the Kolmogorov–Smirnov Z test, and the variables with a normal distribution were expressed as the mean ± standard deviation (SD), while the variables with a nonnormal distribution were expressed as the median [25^th–^75^th^ percentile]. Two groups (with and without hypertension or hyperkinetic versus vascular diseases) were compared for continuous variables via t tests or Mann‒Whitney U tests, as appropriate. Categorical variables were compared via the chi-two test or Fisher’s exact test, as appropriate. Pearson’s correlation coefficients were used to explore the bivariate associations between the examined variables. The other analyses are described in the text. P < 0.05 was considered statistically significant.

## Results

The mean AASI of the population was 0.33 ± 0.17; the median value of the night:day ratio of the number of BP readings was 0.62 [0.55; 0.66].

Table 1 describes the characteristics of the 75 children/adolescents according to the presence of hypertension on the basis of ABPM results. Among the 15 subjects with hypertension, 4 had already received treatment for hypertension (enalapril and/or amlodipine), and 12 had masked hypertension. Compared with the subjects without hypertension, those with hypertension were predominantly male and had increased cf-PWV, decreased baroreflex sensitivity and decreased HRV, while their mean AASI did not differ.

**Table 1.**
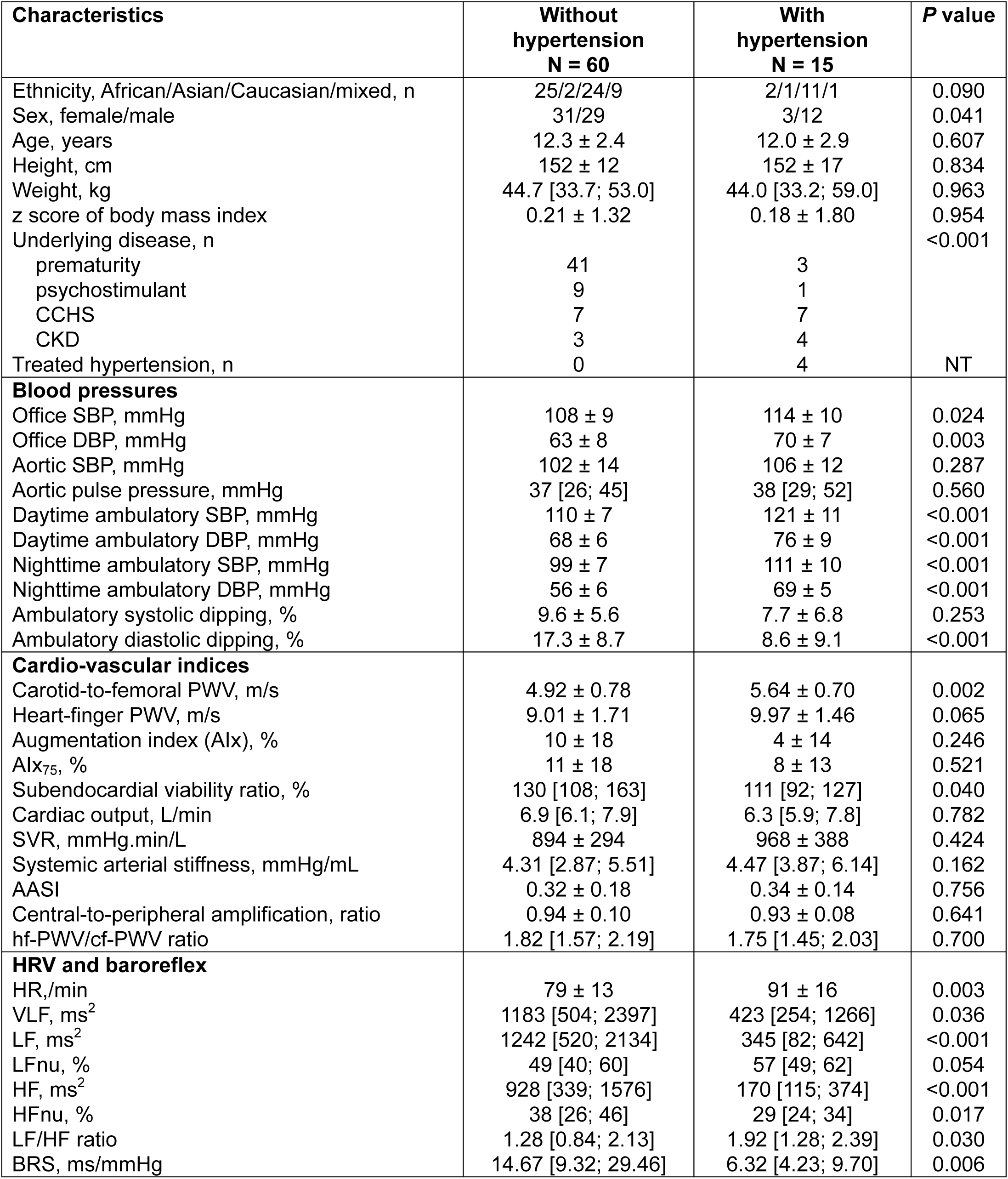
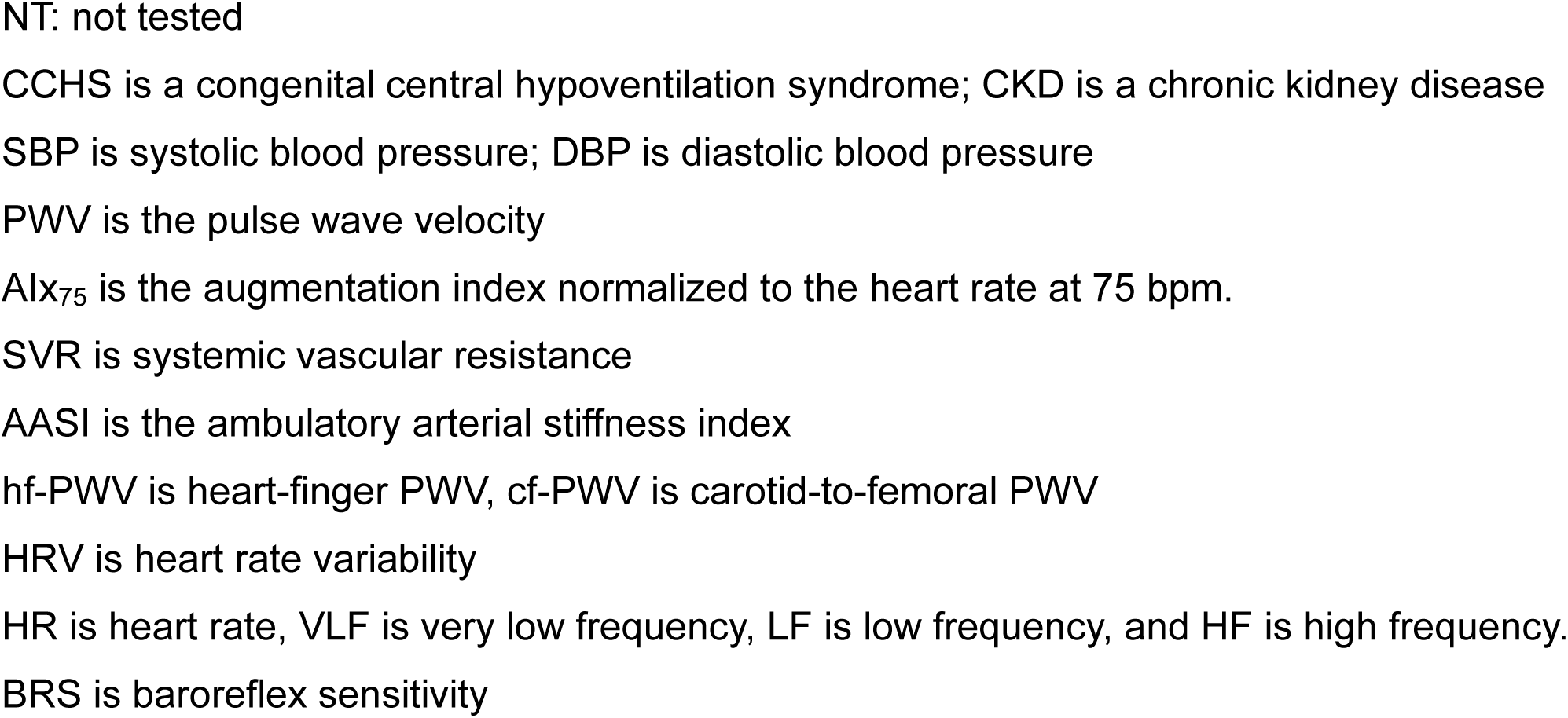
Characteristics of the 75 children/adolescents according to the presence of hypertension.

| Characteristics | Without hypertension<br>N = 60 | With hypertension<br>N = 15 | P value |
| --- | --- | --- | --- |
| Ethnicity, African/Asian/Caucasian/mixed, n | 25/2/24/9 | 2/1/11/1 | 0.090 |
| Sex, female/male | 31/29 | 3/12 | 0.041 |
| Age, years | 12.3 ± 2.4 | 12.0 ± 2.9 | 0.607 |
| Height, cm | 152 ± 12 | 152 ± 17 | 0.834 |
| Weight, kg | 44.7 [33.7; 53.0] | 44.0 [33.2; 59.0] | 0.963 |
| z score of body mass index | 0.21 ± 1.32 | 0.18 ± 1.80 | 0.954 |
| Underlying disease, n |  |  | <0.001 |
| prematurity | 41 | 3 |  |
| psychostimulant | 9 | 1 |  |
| CCHS | 7 | 7 |  |
| CKD | 3 | 4 |  |
| Treated hypertension, n | 0 | 4 | NT |
| **Blood pressures** |  |  |  |
| Office SBP, mmHg | 108 ± 9 | 114 ± 10 | 0.024 |
| Office DBP, mmHg | 63 ± 8 | 70 ± 7 | 0.003 |
| Aortic SBP, mmHg | 102 ± 14 | 106 ± 12 | 0.287 |
| Aortic pulse pressure, mmHg | 37 [26; 45] | 38 [29; 52] | 0.560 |
| Daytime ambulatory SBP, mmHg | 110 ± 7 | 121 ± 11 | <0.001 |
| Daytime ambulatory DBP, mmHg | 68 ± 6 | 76 ± 9 | <0.001 |
| Nighttime ambulatory SBP, mmHg | 99 ± 7 | 111 ± 10 | <0.001 |
| Nighttime ambulatory DBP, mmHg | 56 ± 6 | 69 ± 5 | <0.001 |
| Ambulatory systolic dipping, % | 9.6 ± 5.6 | 7.7 ± 6.8 | 0.253 |
| Ambulatory diastolic dipping, % | 17.3 ± 8.7 | 8.6 ± 9.1 | <0.001 |
| **Cardio-vascular indices** |  |  |  |
| Carotid-to-femoral PWV, m/s | 4.92 ± 0.78 | 5.64 ± 0.70 | 0.002 |
| Heart-finger PWV, m/s | 9.01 ± 1.71 | 9.97 ± 1.46 | 0.065 |
| Augmentation index (Alx), % | 10 ± 18 | 4 ± 14 | 0.246 |
| Alx<sub>75</sub>, % | 11 ± 18 | 8 ± 13 | 0.521 |
| Subendocardial viability ratio, % | 130 [108; 163] | 111 [92; 127] | 0.040 |
| Cardiac output, L/min | 6.9 [6.1; 7.9] | 6.3 [5.9; 7.8] | 0.782 |
| SVR, mmHg.min/L | 894 ± 294 | 968 ± 388 | 0.424 |
| Systemic arterial stiffness, mmHg/mL | 4.31 [2.87; 5.51] | 4.47 [3.87; 6.14] | 0.162 |
| AASI | 0.32 ± 0.18 | 0.34 ± 0.14 | 0.756 |
| Central-to-peripheral amplification, ratio | 0.94 ± 0.10 | 0.93 ± 0.08 | 0.641 |
| hf-PWV/cf-PWV ratio | 1.82 [1.57; 2.19] | 1.75 [1.45; 2.03] | 0.700 |
| **HRV and baroreflex** |  |  |  |
| HR, /min | 79 ± 13 | 91 ± 16 | 0.003 |
| VLF, ms<sup>2</sup> | 1183 [504; 2397] | 423 [254; 1266] | 0.036 |
| LF, ms<sup>2</sup> | 1242 [520; 2134] | 345 [82; 642] | <0.001 |
| LFnu, % | 49 [40; 60] | 57 [49; 62] | 0.054 |
| HF, ms<sup>2</sup> | 928 [339; 1576] | 170 [115; 374] | <0.001 |
| HFnu, % | 38 [26; 46] | 29 [24; 34] | 0.017 |
| LF/HF ratio | 1.28 [0.84; 2.13] | 1.92 [1.28; 2.39] | 0.030 |
| BRS, ms/mmHg | 14.67 [9.32; 29.46] | 6.32 [4.23; 9.70] | 0.006 |

NT: not tested
CCHS is a congenital central hypoventilation syndrome; CKD is a chronic kidney disease
SBP is systolic blood pressure; DBP is diastolic blood pressure
PWV is the pulse wave velocity
Alx<sub>75</sub> is the augmentation index normalized to the heart rate at 75 bpm.
SVR is systemic vascular resistance
AASI is the ambulatory arterial stiffness index
hf-PWV is heart-finger PWV, cf-PWV is carotid-to-femoral PWV
HRV is heart rate variability
HR is heart rate, VLF is very low frequency, LF is low frequency, and HF is high frequency.

Figure 1 shows the correlations of the AASI with different vascular indices and with the nighttime dipping of BP. The AASI did not correlate with either carotid-to-femoral PWV or the AIx in childhood. Additionally, the AASI did not correlate with heart-finger PWV or systemic arterial stiffness (or the two stiffness gradient indices) or with heart rate. AASI did not correlate with carotid-to-femoral PWV in the two subgroups of underlying causes, either vascular (R= 0.06, p=0.648) or hyperkinetic (R= -0.31, p=0.139). In contrast, the AASI was significantly correlated with both systolic and diastolic BP dipping.

**Figure 1.**
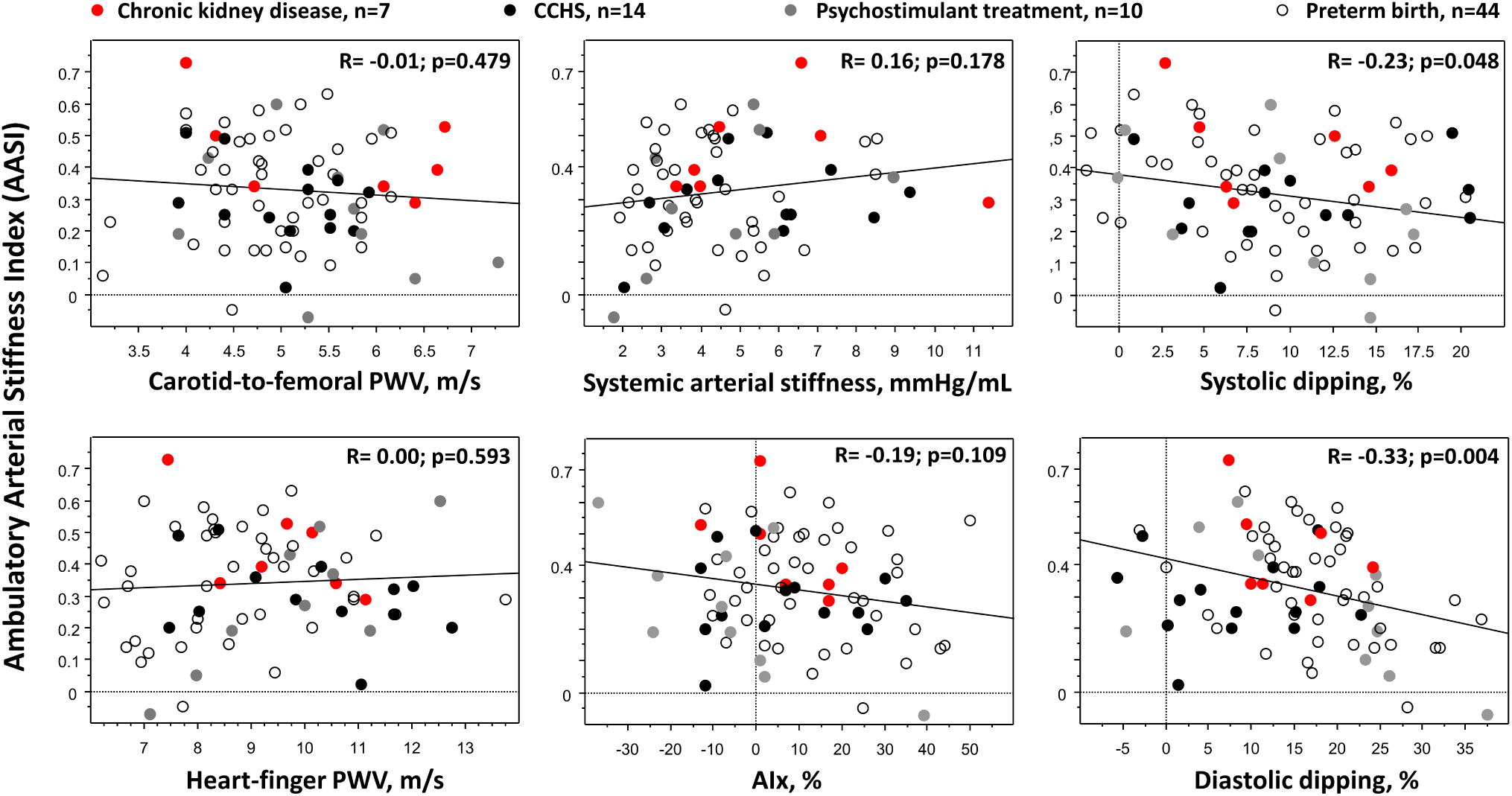
Correlations between the AASI and the other indices. The AASI is described as the dependent variable for four indices of arterial stiffness: carotid-to-femoral pulse wave velocity (PWV), heart-finger PWV, systemic arterial stiffness (aortic pulse pressure/stroke volume) and the AIx (augmentation index). Two additional indices are evaluated: nighttime systolic dipping and diastolic dipping.

We also evaluated the correlates of cf-PWV as described by Schillaci *et al*. ^3^. In univariate analyses, these correlates were age (R= 0.39, p<0.001), office SBP (R= 0.33, p=0.004) and office DBP (R= 0.30, p=0.008) but not supine heart rate (p=0.464).

Given the absence of a significant correlation between the AASI and both cf-PWV and the AIx, we evaluated whether the different vascular indices obtained were correlated, suggesting their validity. In young normotensive subjects, the stiffness gradient is illustrated by the increase in arterial stiffness from upstream proximal large arteries to downstream distal medium-sized arteries. We first evaluated whether the two calculated indices of the stiffness gradient were correlated: the central-to-peripheral amplification ratio was correlated with hf-PWV/cf-PWV, R= 0.31; p=0.009. Systemic arterial stiffness also correlated with these two calculated gradients: the central-to-peripheral amplification ratio (R= 0.46, p<0.001) and the hf-PWV/cf-PWV (R= 0.25; p=0.047).

Systemic arterial stiffness has previously been evaluated in hypertensive adults via M-mode echocardiography ^21^. Since we used a different method to evaluate stroke volume in a pediatric population, we evaluated its correlations with other vascular indices, which are described in Figure 2. The figure shows that this arterial stiffness index was correlated with heart-finger PWV and the AIx_75_ (and with the AIx: R= -0.31, p=0.009). Aortic SBP and baroreflex are also correlated with systemic arterial stiffness.

**Figure 2.**
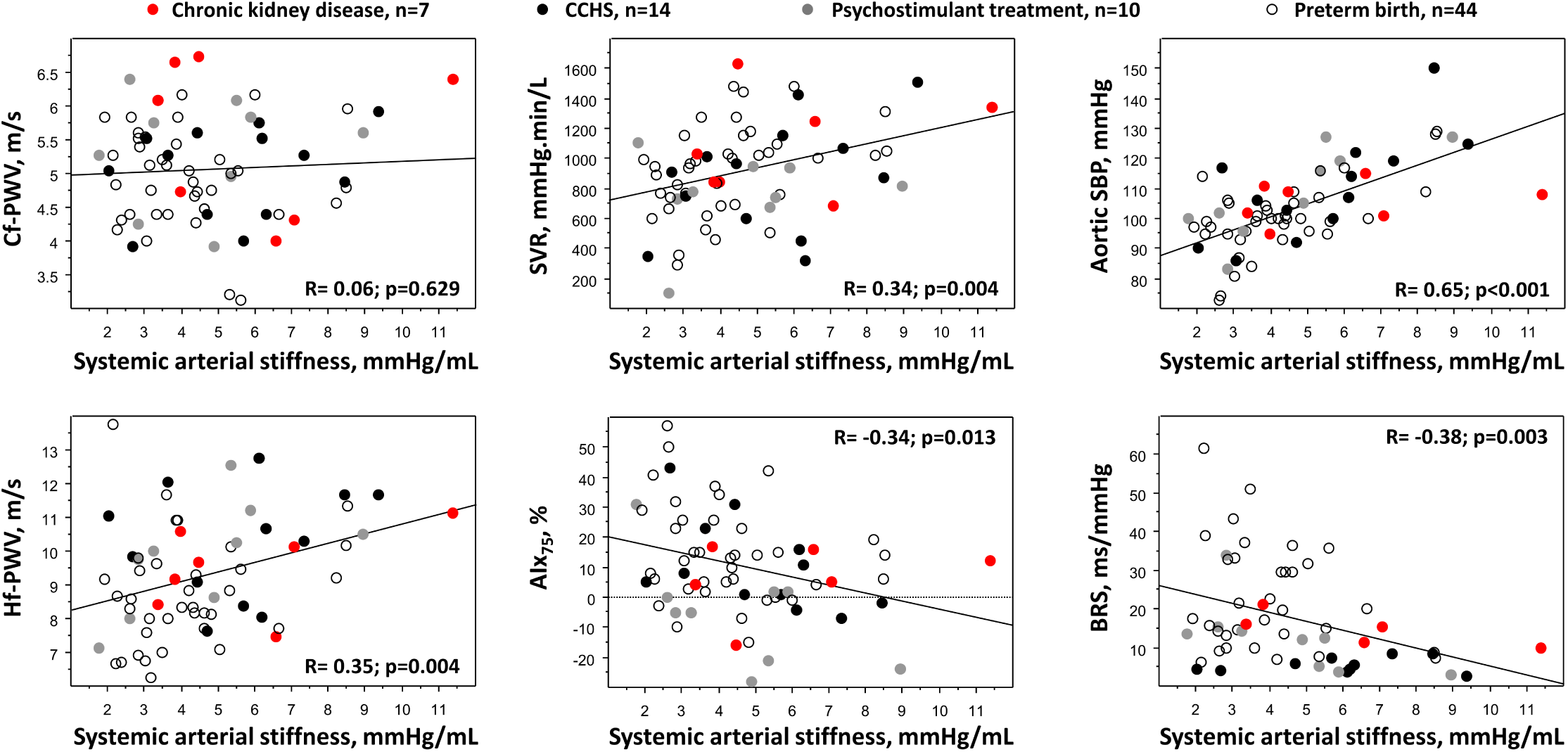
Evaluation of the correlates of systemic arterial stiffness. Systemic arterial stiffness (aortic pulse pressure/stroke volume) is described as an independent variable for four indices of arterial stiffness (carotid-to-femoral pulse wave velocity (cf-PWV), heart-finger PWV (hf-PWV), systemic vascular resistance (SVR) and AIx_75_ (augmentation index, normalized at 75 bpm)). Two additional indices are evaluated: aortic systolic blood pressure (SBP) and baroreflex sensitivity (BRS). The latter index was obtained for 61/75 participants (in some subjects, ascendant or descendant BRSs could not be computed because of either failure to obtain the required criteria for computation or because the device slipped out of position while the subject was standing).

In older subjects, aging-related vascular remodeling is characterized by an increase in the AIx (more positive values). The negative correlation between arterial stiffness and the AIx_75_ may seem surprising. We thus evaluated the correlates of AIx_75_. AIx_75_ was correlated with height (R= -0.27; p=0.024), aortic SBP (R= -0.24; p=0.040), the central-to-peripheral amplification ratio (R= -0.31; p=0.007) and aortic pulse pressure (R= -0.32; p=0.005). The AIx_75_ did not significantly correlate with age (p=0.166), office DBP (p=0.555), daytime ABPM-DBP (p=0.081) or cf-PWV (p=0.707). In a multiple regression analysis with AIx_75_ as the dependent variable and height and the central-to-peripheral amplification ratio as the independent variables (the other variables were not included due to collinearity), these two latter variables remained independently associated with AIx_75_ (adjusted r^2^ of the model = 0.13, p = 0.003).

Finally, we compared the characteristics of the children/adolescents according to the underlying condition, namely, vascular disease (preterm birth and CKD, n=51) versus hyperkinetic disease (CCHS and psychostimulant treatment, n=24), as shown in Table 2. Participants with hyperkinetic diseases were characterized by a higher heart rate, higher cardiac output, lower HRV and lower diastolic BP dipping, resulting in higher nighttime DBP. Their mean aortic SBP and heart-finger PWV were greater than those of participants with vascular disease.

**Table 2.** Characteristic according to the two kinds of disease, either vascular or hyperkinetic.

| Characteristics | Vascular diseases<br>N = 51 | Hyperkinetic diseases<br>N = 24 | P value |
| --- | --- | --- | --- |
| Ethnicity, African/Asian/Caucasian/mixed, n | 20/3/18/10 | 7/0/17/0 | 0.012 |
| Sex, female/male | 22/29 | 12/12 | 0.578 |
| Age, years | 12.1 ± 2.2 | 12.6 ± 3.1 | 0.469 |
| Height, cm | 151 ± 12 | 153 ± 15 | 0.611 |
| Hypertension, n | 7 | 8 | 0.065 |
| Treated hypertension, n | 4 | 0 | NT |
| **Blood pressures** |  |  |  |
| Office SBP, mmHg | 108 ± 9 | 111 ± 10 | 0.175 |
| Office DBP, mmHg | 63 ± 7 | 67 ± 10 | 0.075 |
| Aortic SBP, mmHg | 100 ± 11 | 108 ± 16 | 0.012 |
| Aortic pulse pressure, mmHg | 37 [25; 44] | 38 [27; 53] | 0.532 |
| Daytime ambulatory SBP, mmHg | 112 ± 9 | 114 ± 8 | 0.263 |
| Daytime ambulatory DBP, mmHg | 69 ± 7 | 71 ± 7 | 0.309 |
| Nighttime ambulatory SBP, mmHg | 102 ± 10 | 102 ± 6 | 0.738 |
| Nighttime ambulatory DBP, mmHg | 57 ± 7 | 62 ± 7 | 0.008 |
| Ambulatory systolic dipping, % | 8.9 ± 5.7 | 10.0 ± 6.3 | 0.461 |
| Ambulatory diastolic dipping, % | 17.2 ± 8.0 | 12.3 ± 11.4 | 0.035 |
| **Cardio-vascular indices** |  |  |  |
| Carotid-to-femoral PWV, m/s | 4.98 ± 0.80 | 5.25 ± 0.83 | 0.181 |
| Heart-finger PWV, m/s | 8.85 ± 1.60 | 9.94 ± 1.70 | 0.012 |
| Augmentation index (Alx), % | 12 ± 16 | 1 ± 19 | 0.013 |
| Alx<sub>75</sub>, % | 13 ± 16 | 4 ± 18 | 0.035 |
| Subendocardial viability ratio, % | 125 [108; 152] | 121 [95; 146] | 0.366 |
| Cardiac output, L/min | 6.4 [5.5; 7.5] | 7.6 [6.2; 8.2] | 0.048 |
| SVR, mmHg.min/L | 831 ± 330 | 949 ± 303 | 0.140 |
| Systemic arterial stiffness, mmHg/mL | 4.00 [3.05; 5.33] | 5.12 [3.05; 6.18] | 0.208 |
| AASI | 0.35 ± 0.17 | 0.28 ± 0.16 | 0.095 |
| Central-to-peripheral amplification, ratio | 0.92 ± 0.07 | 0.97 ± 0.12 | 0.031 |
| hf-PWV/cf-PWV ratio | 1.75 [1.51; 2.05] | 1.96 [1.69; 2.28] | 0.129 |
| **HRV and baroreflex** |  |  |  |
| HR, /min | 78 ± 13 | 88 ± 15 | 0.006 |
| VLF, ms<sup>2</sup> | 1393 [660; 2680] | 542 [277; 849] | 0.002 |
| LF, ms<sup>2</sup> | 1309 [522; 2134] | 385 [164; 953] | 0.002 |
| LFnu, % | 49 [39; 60] | 57 [47; 61] | 0.041 |
| HF, ms<sup>2</sup> | 1010 [536; 1841] | 170 [116; 522] | <0.001 |
| HFnu, % | 40 [27; 46] | 30 [24; 35] | 0.004 |
| LF/HF ratio | 1.18 [0.80; 1.98] | 1.94 [1.32; 2.42] | 0.005 |
| BRS, ms/mmHg | 16.46 [9.99; 31.84] | 5.74 [3.98; 12.38] | <0.001 |

See Table 1 legend for abbreviations.

## Discussion

The main result of our cross-sectional study, which used different methods for the assessment of vascular remodeling, was that the AASI is not a reliable marker of arterial stiffness in childhood.

The first issue addresses methodological aspects. We used the same devices that were used in the studies of Li *et al*.^2^ and Schillaci *et al*.^3^ for AASI calculations (Spacelabs model) and cf-PWV measurements (SphygmoCor system). Nevertheless, AIx_75_ was not obtained from a transfer function of the radial waveform but from the carotid waveform, which may be a strength given the criticisms made to transfer function.^7^ We further calculated systemic arterial stiffness (aortic pulse pressure/stroke volume) and measured heart-finger PWV as an additional and complementary means of vascular structure assessment. As the distance from the heart increases, the elastin content of the arterial structure decreases, the collagen fiber content increases, and the diameter decreases, resulting in increased stiffness of the aorta to the peripheral arteries. The heart-finger PWV combines the elasticity of the central aorta, middle arteries, small arteries and microvessels.^18^ The magnitude of systemic arterial stiffness was in line with that measured in healthy subjects^22^ and correlated with the stiffness of peripheral arteries, i.e., heart-finger PWV and AIx_75_, and with central indices, such as aortic SBP and baroreflex sensitivity. The two stiffness gradient indices (hf-PWV/cf-PWV and the central-to-peripheral amplification ratio) were correlated to some extent. Overall, the correlations observed via different methods of assessment suggest the validity of our measurements.

The main result is the absence of a significant correlation between the AASI and cf-PWV, both in the whole population and in the two subgroups of underlying conditions, which is at variance with that observed in adulthood.^2^ Schillaci *et al*. further suggested that the observed correlation could be related to confounders since the relationship between the AASI and cf-PWV was no longer significant in a multivariate regression analysis model.^3^ Our results in childhood favor the latter results. In a recent review dealing with arterial pulse wave modeling, the authors stated that the AASI is more a measure of ventriculo-arterial coupling determined by heart rate and vascular resistance.^23^

We found that the AASI was correlated with both diastolic and systolic dipping, and the correlation coefficients were quite similar to those previously reported.^3,24^ We confirmed that the AASI is inversely correlated with the nocturnal decrease in blood pressure, especially in ambulatory recordings with a disproportionately large number of nighttime readings (62% in our series), as previously suggested.^24^ These positive results further support the validity of our negative results concerning the AASI and arterial stiffness.

In our hypertensive subjects, we observed a significant increase in carotid-to-femoral PWV. Earlier aortic aging in hypertension patients is counterbalanced by a significant BP-related upward reset of peripheral muscular artery PWV (only a trend was observed for heart-finger PWV in our study).^25^ In larger conductive arteries, vessel remodeling in hypertension leads to increased vessel stiffness, which could negatively impact myocardial work capacity and coronary perfusion, as suggested by a lower subendocardial viability ratio in our hypertensive subjects. Finally, reduced HRV is also a well-known characteristic of hypertension, occurring even before the BP increases.^26^ Thus, the characteristics of our subjects with hypertension were as expected, suggesting that the absence of an increase in the AASI in our study is a true finding, which is the opposite of that reported by Simonetti *et al*. in children.^5^ Nevertheless, this opposite result may also be related to the larger sample size in the study of Simonetti *et al*. and the different causes of hypertension in their study (essential hypertension, 51% and CKD, 37%).^5^

In younger subjects, high SBP at the brachial artery may not reflect high arterial stiffness since it overestimates central aortic pressure values (amplification of the pressure waves) and is often related to sympathetic overactivity (high cardiac output).^27^ Similarly, the aortic SBP was not greater in our hypertensive subjects than in those without hypertension, suggesting that some children had spurious hypertension.^28^ The fact that the AASI is associated with arterial parameters even in younger subjects makes the AASI an interesting index for the evaluation of arterial alterations in younger subjects,^2,29^ but our results contradict this hypothesis, which deserves to be demonstrated.

Systemic arterial stiffness and AIx_75_ were negatively correlated, suggesting that negative values of AIx_75_ are associated with increased stiffness, which may seem counterintuitive since AIx, which is supposed to represent the relative contribution of the reflected wave to central pulse pressure, increases from young adulthood to midlife. Nevertheless, AIx_75_ has been shown to decrease from 2 to ∼14 years of age in healthy children.^30,31^ In our study, the AIx_75_ was negatively correlated with height in childhood, as previously reported in both adulthood^32^ and childhood,^30,33^ suggesting the validity of our results. The AIx is defined as the difference between the second and first systolic peaks of the central arterial waveform, expressed as a percentage of the central pulse pressure, and negative values of the AIx are encountered in young individuals. We found a negative correlation between AIx_75_ and the central-to-peripheral amplification ratio: when the central SBP was greater than the peripheral SBP (ratio > 1, inverse stiffness gradient), it was associated with negative values of AIx_75_. Thus, AIx_75_ is not an index of reflected waves in youth, as previously demonstrated by Hughes *et al*.^34^ In the case of waveforms with negative values of AIx, problems of interpretation are confounded by the presence of a forward traveling decompression wave that causes a late shoulder in the pressure waveform and gives rise to negative values of AIx. The mechanism accounting for the forward decompression wave in mid-systole remains to be fully established.^34^ Overall, the assumption that a higher AIx represents the integrated effects of aortic stiffening, increased PWV and earlier return of a potentially larger backward wave may be overly simplistic, as previously stated.^32^ Numerous modeling studies have been conducted to explain these waveforms, emphasizing the complexity of their interpretation.^23^ Overall, a more negative value of AIx_75_ may represent an adaptive mechanism to increase systemic stiffness, decreasing peripheral SBP compared with central SBP, as previously suggested.^35^

The comparison of the subjects according to the king of underlying disease given in Table 2 may seem to be an oversimplification since most participants had normal cardiovascular function. Nevertheless, hyperkinetic and vascular causes can be effectively differentiated, and hyperkinetic causes are associated with vasoconstriction due to increased sympathetic tone, leading to an increase in heart-finger PWV and a decrease in HRV.^36^ Interestingly, both AIx and AIX75 were lower in subjects with hyperkinetic conditions than in those with vascular conditions, which is in line with the decrease in AIx in obese children compared with healthy children with normal weight,^35^ since obesity is also a hyperkinetic condition.^37^

Our study has at least one limitation related to our design, including only secondary causes of hypertension, which limits the generalizability of our findings. We selected subjects with four different conditions, two associated with neurogenic hypertension and two with vascular hypertension, since these different mechanisms may have affected the correlations. Figures 1 and 2 suggest that our results were valid regardless of the underlying cause.

In conclusion, our cross-sectional study revealed that the AASI is not related to any marker of arterial stiffness in childhood and cannot constitute an interesting index for the evaluation of arterial alterations in younger subjects.

## Data Availability

All data produced in the present study are available upon reasonable request to the authors

